# Evaluating user-operated tests of audible contrast sensitivity towards efficient hearing healthcare services

**DOI:** 10.1101/2025.07.28.25332303

**Authors:** Raul Sanchez-Lopez, Johannes Zaar, Søren Laugesen

## Abstract

The user-operated audiometry (UAud) project aims at introducing an automated system for user-operated audiometric testing into everyday clinical practice. Here, we focus on the Audible Contrast Threshold (ACT™) test, which has been proposed as a language-independent alternative to aided speech-in-noise tests. Five distinct user-operated ACT (U-ACT) test candidates were evaluated in terms of performance, reliability, and usability against two established test benchmarks. The study involved 28 participants with diverse hearing and cognitive abilities. The results demonstrated that the primary factor influencing the reliability of test results was the type of *task*. In terms of usability, the participants reported positive experience with all test candidates, with the Yes/No task resulting in the lowest perceived difficulty and best understanding. Overall, a test candidate with a custom adaptive procedure inspired by audiologists’ experiences with ACT was chosen for the UAud protocol as a user-operated test of hearing-in-noise perception.

## Introduction

The total number of people living with some degree of hearing loss worldwide is projected to rise from approximately 1.57 billion in 2019 to 2.45 billion by 2050. This represents an increase of 56.1% more individuals who will need hearing healthcare services^1^.

Furthermore, it is expected that more than 44% of individuals aged 70 years and older will experience hearing deficits. With this perspective, and an increasing workforce shortage in hearing healthcare^2^, there exists an urgent need to improve efficiency and reduce costs in hearing healthcare to ensure access to quality services^1,3,4^. The currently used hearing tests require active participation of a hearing care professional, and represent a bottleneck that effectively limits access to hearing healthcare^5,6^. To meet the demand for hearing assessments, the availability of well-trained audiologists and clinical facilities is crucial^7^.

However, audiologists are responsible for various important tasks beyond hearing diagnostic tests such as hearing-aid fitting and verification as well as counselling^8^. The User-operated Audiometry (UAud) project seeks to introduce an automated system that enables user-operated audiometric testing within everyday clinical practice, aiming to support audiologists in prioritizing hearing healthcare services and thereby allowing them to treat more patients and provide better hearing rehabilitation^9^.

The most prevalent form of auditory impairment is presbycusis, or age-related hearing loss, which typically affects individuals over the age of 55^10^. This progressive condition significantly impacts communication and overall quality of life. Notably, speech perception abilities are closely linked to cognitive factors such as working memory^11–13^. Furthermore, cognitive decline is strongly associated with auditory deprivation; unaddressed hearing loss has been identified as the leading modifiable risk factor for dementia^14,15^. Within the scope of the UAud project, individuals with comorbid hearing loss and cognitive decline may encounter difficulties with user-operated testing. Elderly people with cognitive impairment generally require more time and more assistance to complete tasks compared to healthy patients when performing behavioral tests or using technology in general ^16,17^. It is, therefore, essential to design automated protocols that are accessible to a broad population, regardless of their cognitive profile^18,19^.

Currently, audiological assessments are mainly based on pure-tone audiometry, which consists of the detection of pure tones played at different frequencies and sound pressure levels to obtain the patient’s hearing thresholds^8^. However, there has been a growing focus on additionally evaluating supra-threshold abilities^11,20^, such as the patient’s ability to discriminate speech in the presence of noise when the speech is fully audible^21^. The clinical evaluation of realistic, or ecologically valid, aided speech-in-noise perception presents several challenges, such as the need for multi-loudspeaker setups and speech material in multiple languages^22,23^. This has motivated the search for alternative tests capable of capturing the fundamental abilities employed in challenging ‘cocktail-party’ scenarios, which are characterized by multiple interfering sound sources including speech^24–26^. However, despite decades of research, the clinical characterization of hearing deficits beyond elevated hearing thresholds remains a challenge.

One promising approach involves the detection of spectro-temporal modulations (STM) imposed on broadband sounds^26–28^. These modulations create spectral and temporal ripples that mimic the rapid fluctuations of natural speech. A key advantage of STM stimuli is the precise control of modulation levels within classical psychoacoustic adaptive procedures^28,29^. This paradigm is analogous to contrast sensitivity testing in the visual domain, which utilizes spatial and temporal patterns^30,31^. Because the ability to discriminate subtle cues in noisy speech is strongly associated with STM sensitivity^32,33^, these tests have been proposed both as a proxy for speech-in-noise perception in ecologically valid environments and as a predictor of hearing-aid benefit^34^.

Recently, the Audible Contrast Threshold (ACT™) test^35^ was introduced as a quick, simple, and clinically viable manual version of an STM sensitivity test^28,33,34^. The results of ACT and its precursor, the STM test^34^, showed high correlations with speech reception thresholds measured in hearing-in-noise tests. The speech reception threshold is defined as the speech sound level, relative to the interfering noise, at which an individual can repeat 50% of the speech material (sentences in this case) in an adaptive test. Notably, this correlation was stronger when using realistic speech-in-noise conditions with spatially distributed speech interferers^28^. Although the addition of ACT in current clinical practice is less resource-demanding than other time-consuming audiometric tests, an automated implementation of ACT offers significant benefits for busy clinical services, making it a practical choice alongside the automatic method for testing auditory sensitivity (AMTAS)^6,36^.

In the present study, we therefore evaluated distinct candidate user-operated tests of audible contrast sensitivity (U-ACT), which assess the supra-threshold hearing-in-noise abilities of the patient with language-independent stimuli and can thus complement the pure-tone audiometric testing. Both the ACT and the STM tests were considered here as test *benchmarks,* to which the different U-ACT test candidates were compared. While ACT is a test administered by an audiologist, STM is a user-operated test conducted using a research platform for psychophysical experiments. For the U-ACT tests we considered different *tasks* and *procedures*. In this context, a *task* is the type of patient action that produces a response in the test (e.g., pressing a button upon detecting an event), while a *procedure* is the method used to seek the threshold, for example, decreasing the contrast level (the strength of the modulation) by a certain value every time there is a response until no further responses are obtained.

The main goal of the present study was to identify an optimal candidate for a user-operated ACT test that is clinically feasible and adheres to the stated objectives. The objectives for a successful user-operated test were: 1) The test should be easy to perform such that the patient immediately understands what needs to be done. 2) The test should not be unnecessarily long; it must allow patients to maintain attention and performance effectively. 3) The test must be reliable and repeatable, demonstrating good agreement with standard or well-accepted alternatives (benchmarks). 4) Overall, the test should be accessible to a broad group of patients without requiring any special modifications for subpopulations, including those with low cognitive abilities. 5) The test should have an optimal quality of interaction so the patients can perform the test seamlessly. In the present study, we proposed a user-centered multi-dimensional evaluation which aims at providing evidence of the feasibility and adequacy of the new user-operated test in a realistic case scenario and with a diverse group of participants.

## Experimental design and study framework

### Multidimensional user-centered evaluation

The study design was inspired by the Roles Activities Materials Environment System (RAMES) framework which supports user-centered evaluations in research studies^37^. The objective of the framework is to assess all the factors affecting the outcomes of the evaluation, knowing the interaction among the different elements. Additionally, the design was guided by the ISO/IEC 25022:2016^38^ standard considering the quality in use with focus on the context coverage. The context coverage is related to efficacy, efficiency, and satisfaction and it is defined as the extent to which the system can be used by people without specific knowledge, skills, or experience. The definition of best candidate was based on the criteria summarized in Table 1.

**Table 1:** Criteria for the definition of best U-ACT test candidate.

| Psychoacoustics | User interaction | Audiometric groups | Cognition groups |
| --- | --- | --- | --- |
| High Reliability<br>Low training effect | High success rate<br>Low time on task<br>Low self-reported<br>Difficulty | All groups can<br>perform the test | All groups can<br>perform the test |

### Study participants

The study sample comprised 24 hearing-impaired (HI) participants with diverse hearing and cognitive abilities, all of them above 50 years old, along with 6 young normal-hearing (NH) listeners aged below 30 years. The HI participants were divided into two groups according to their cognitive abilities. To assess the audiological characteristics, the participants underwent standard pure-tone audiometry as part of the protocol.

Furthermore, their cognitive abilities were assessed using the reversed digit span^39,40^ (RDS) test, which specifically evaluates the working memory performance of the participant.

### Study protocol

The study protocol involved two *visits*, each comprising two *blocks*. Within each *block*, participants completed one run of each of five user-operated ACT test candidates, presented in random order. Data from all four *blocks* were utilized for various investigations into the feasibility of the test candidates. *Block* 1 evaluated instruction efficacy, blocks 2 & 4 assessed measurement reliability and quality, while block 3 provided a fresh run after one week. This fresh run was intended to simulate a follow-up visit where the test is repeated but it is not the first time that the participant experiences the test.

In the present study, we specifically focused on exploring the utility of language-independent instructions in user-operated tests. To achieve this, we implemented a set of non-verbal instructions consisting of four main steps: 1) putting the headphones on, 2) familiarization, 3) training, and 4) demonstration. These instructions were refined through multiple iterations with naïve users interacting with the system^41^ who were not part of the final study sample. During this pre-investigation, we identified key graphical signifiers (a set of icons), that facilitated correct instruction usage while allowing for user mistakes.

After the pre-investigations, we defined efficacy based on a primary success rate reflecting whether the threshold obtained in *block 1* was comparable to the one obtained in the remaining *blocks*. Additionally, we monitored the frequency of support interventions by tracking participant requests for written instructions, supplementary visual cues, or examiner assistance.

Within the two visits, two benchmark tests of the participants’ ability to detect spectro-temporal modulations were used: (i) a research version (STM) ^34^ and (ii) the clinically viable ACT test^35^. STM is user-operated and uses a 3-alternative forced choice task and a transformed up-down adaptive procedure^29^, while ACT is administered by an audiologist. The ACT test consists of a running stimulus of consecutive 1-second sound segments.

The audiologist activates the target stimulus which adds a spectro-temporal modulation with a specific contrast level to the sound, and follows a Hughson-Westlake procedure (similar to the pure-tone audiometry)^42^ to seek the threshold of the patient based on their responses. The tests benchmarks were tested between *blocks* 1 and 2 in *visit 1,* and between *blocks* 3 and 4 in *visit 2*.

Participants completed a subjective evaluation immediately after each test (except for the initial *block*). The evaluation consisted of a series of statements rated using a continuous visual analog scale (VAS)^43^ from “completely disagree” to “completely agree”. The statements were divided into two categories: test attributes and self-confidence with the test. Test attributes included difficulty, perceived length, demand, and boredom. Self-confidence questions aimed at determining if participants understood the instructions, found the test useful, thought that the results were good, and what their overall opinion of the test was. To ensure the reliability of the subjective usability metrics, the experimental design utilized both positive and negative linguistic framing for the attributes (*wording*).

This approach was implemented to control and assess the stability of participants’ ratings.

### User-operated Audible Contrast Threshold test candidates

We designed and implemented a total of five U-ACT test candidates which differed in terms of *task* and *procedure*. In a non-automatic test, the *task* is related to the instructions given to the participant while the *procedure* consists of step-by-step actions followed by the examiner. In user-operated tests, the *procedure* is automated. Each U-ACT candidate was numbered from 0 (U-ACT-0) through 4 (U-ACT-4). U-ACT-0 mirrors the standard ACT test by using the same *task* and *procedure* (but in an automatic manner), while U-ACT-4 aligns closely with the STM test’s *task* and *procedure*. The present multidimensional evaluation aims to compare three tests termed U-ACT-1, U-ACT-2, and U-ACT-3, sharing a custom psychoacoustic procedure but differing in *task* configuration. These tests are evaluated alongside U-ACT-0 and U-ACT-4, which closely resemble the established benchmarks.

Figure 1 depicts the five U-ACT candidates showing their progression from quick-and-simple ACT-like (U-ACT-0, blue background), to the research-oriented STM-like (U-ACT-4, orange background) designs. The three intermediate candidates (green background) differ only in their task configuration. Note that the 1-button task is only compatible with a continuous stimulus. In terms of the stimulus presentation, U-ACT-1 and U-ACT-0 used a “running stimulus” paradigm requiring responses ‘when’ the target was presented. As depicted in Figure 1, the user interface consisted of a single response button. The test candidates U-ACT-2, U-ACT-3, and U-ACT-4 employed a sequential presentation of three stimuli with brief pauses between intervals. The sequential presentation tests differed in terms of *task*: in U-ACT-2 the first and third intervals were always references, while the second interval contained either a target or a (catch-trial) reference (a ‘which’ *task*) and the user interface accordingly consisted of two buttons representing a target (left button) and a reference (right button). U-ACT-3 and U-ACT-4 applied a 3-Alternative Forced Choice (3-AFC) *task* (a ‘where’ task) where the target is in one of the three intervals and the user interface consists of three buttons corresponding to the three intervals. In terms of *procedure*, U-ACT-1, U-ACT-2 and U-ACT-3 share a Bayesian-based custom tracking *procedure* (see Methods section) instead of typical up-down adaptive procedures. The *procedure* was inspired by the experience of audiologists with the manual ACT, and it utilizes a Bayesian approach to decrease the uncertainty of the measure (the participant’s threshold) with each new stimulus presentation.

**Figure 1:**
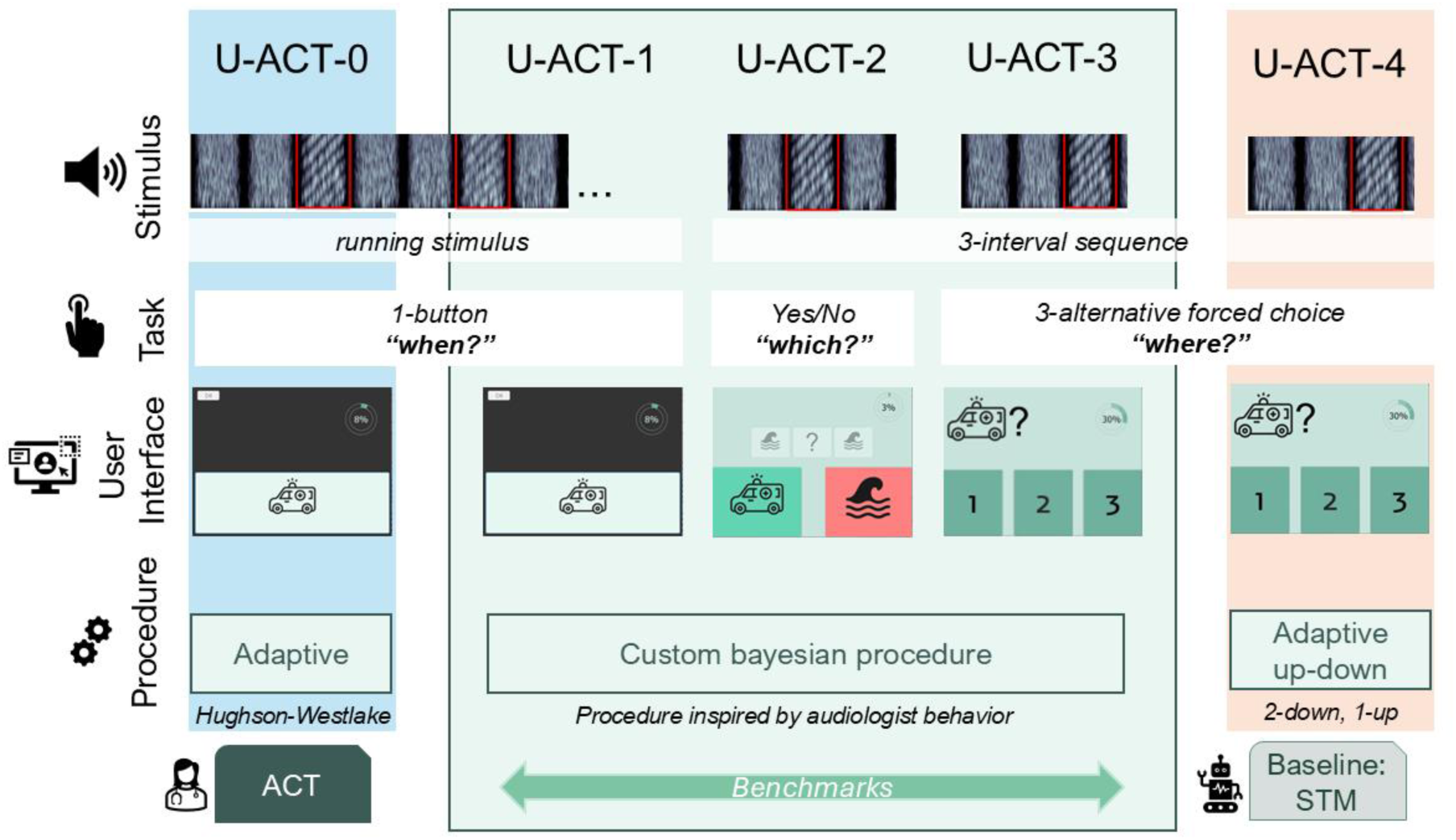
Illustration of the essential aspects of the user-operated Audible Contrast Threshold (U-ACT) test candidates, in terms of stimulus, task, user interface, procedure, and similarity with the test benchmarks. In the user-operated domain, we introduced a visual communication system in which the target stimulus is referred to as an’ambulance’, and the reference sound is described as a’wave’. From left to right, the test candidates are designated with an increasing number related to their resemblance with the clinical Audible Contrast Threshold (ACT) test and spectro-temporal modulation detection (STM) test.

## Results

### Participants’ hearing and cognitive abilities

**Figure 2Error! Reference source not found.** shows the results from pure-tone audiometric assessments, cognitive assessments based on RDS scores, and participants’ ages. The HI participants were divided into two groups based on their RDS scores: a group of 11 people with nominally higher cognitive abilities (HI_hi_) and a group of 11 with lower cognitive abilities (HI_lo_). This categorization is visually represented by a dashed line in the corresponding panel (third panel from the left) of Figure 2**Error! Reference source not found..**

**Figure 2:**
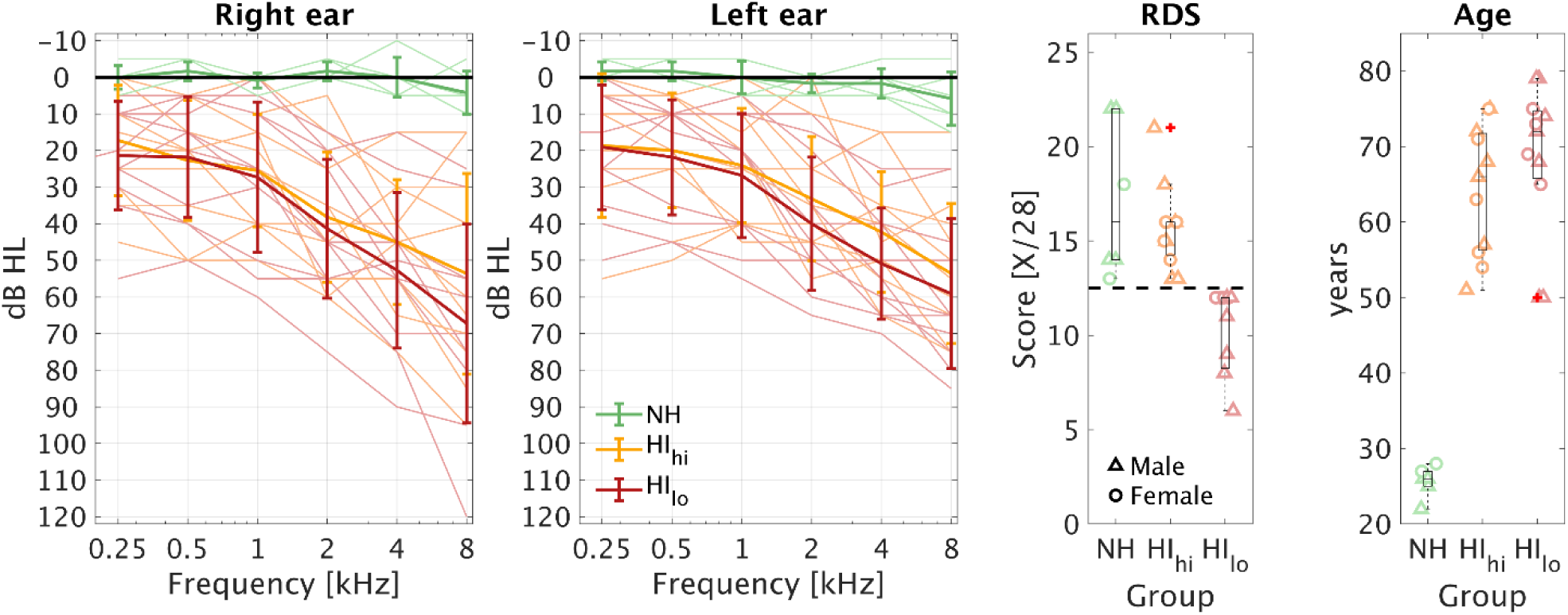
Summary of the participant characteristics. First and second panels from the left represent the average pure-tone thresholds (bold lines) with maximum and minimums (whiskers) and raw data (thin lines) corresponding to each of the three participant groups (NH: Normal hearing in green, HI_hi_: Hearing-impaired with higher cognitive abilities in orange, HI_lo_: Hearing-impaired with lower cognitive abilities in red). Third panel from the left: Reverse Digit Span (RDS) scores, which were the basis for splitting the hearing-impaired group into HI_hi_ and HI_lo_ (separation indicated by black dashed line). Rightmost panel: Age, for the three participant groups. boxes indicate the 25th, 50th (median), and 75th percentiles; whiskers represent the range of non-outlier data, and symbols (+) denote values exceeding 1.5 times the interquartile range…

The average hearing thresholds among participants with hearing loss showed no noteworthy differences between the two HI groups, with average thresholds below 1 kHz of about 20 dB hearing level (HL) and a gradual increase in thresholds at higher frequencies (between 40 and 60 dB HL). On average, the hearing thresholds were slightly lower in the left ear than in the right ear in HI listeners. This was an incidental finding, as there were no specific symmetry requirements for audiograms during recruitment. The NH group scored between 14 and 22 on the RDS (maximum score of 28), averaging approximately 17.8 points. The participants with hearing loss and higher cognitive abilities scored between 14 and 21 whereas the ones with lower cognitive abilities scored between 6 and 12. In terms of age, the HI_lo_ group was slightly older (median = 72 years) than the HI_hi_ group (median = 66 years).

### Test benchmarks are comparable

The results from the two benchmarks across all participants are displayed and compared in Figure 3.

**Figure 3:**
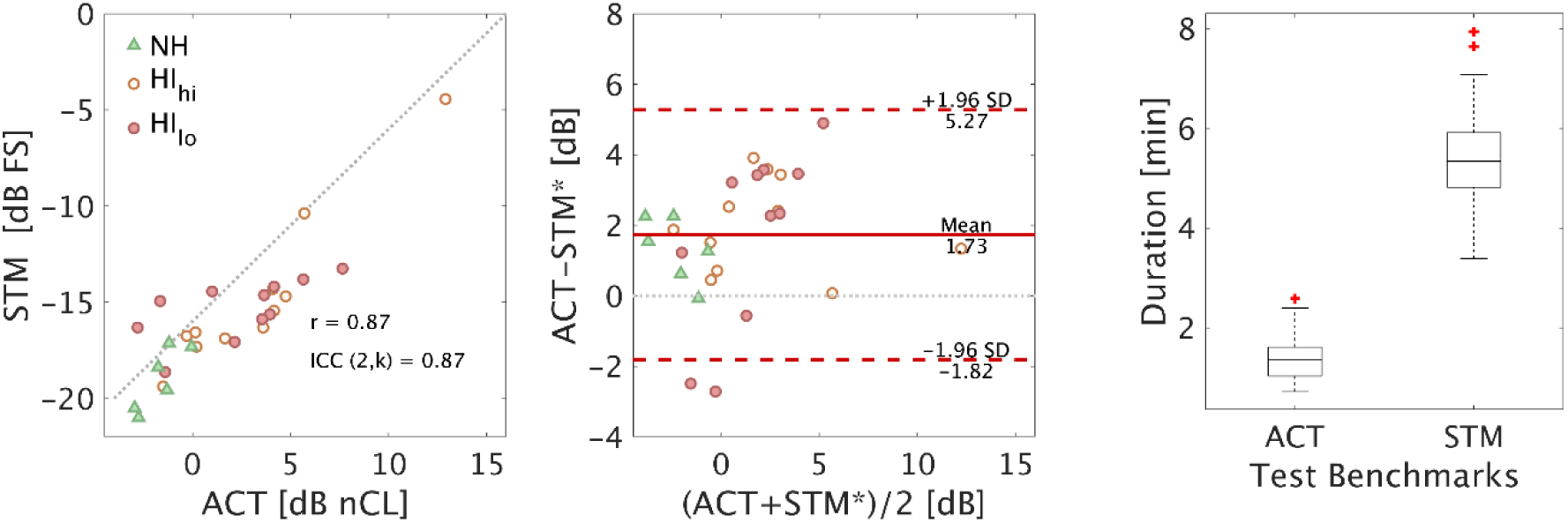
Comparison between the spectro-temporal modulation detection (STM) test and the Audible Contrast Threshold (ACT) test. Left panel: Scatter plot of agreement between tests. Middle panel: Bland-Altman plot of differences between tests. STM* denotes that the thresholds have been transformed in dB nCL for better comparison with ACT. Right panel: Differences in duration, boxes represent the interquartile difference and whiskers the maximum and minimum values not considering outliers. Red crosses indicate outliers.

The left panel shows the STM results as a function of the ACT results. ACT (and the result of the U-ACT variants) is measured in normalized contrast level (nCL), a quantity that maps the physical magnitude (modulation level) to perceived contrast. Thus, an ACT score of 0 dB nCL represents the median performance in listeners with normal hearing^35^ and an elevated threshold indicates a contrast loss. In contrast, the STM test result is expressed in modulation depth in dB full scale (FS), where 0 dB FS corresponds to fully modulated and it is equivalent to 16 dB nCL^35^. For the Bland-Altman analysis, the dB-FS results from STM were transformed to dB nCL by adding 16 dB. The intraclass correlation coefficient between the STM and ACT test results was very high (ICC(2,k) = 0.87) indicating excellent agreement. The participants’ thresholds spanned a range of-2.95 to 12.9 dB nCL, with a mean value of 1.8 dB nCL. The differences, depicted in the middle panel, were smaller than ±2 dB with a bias of 1.73 dB showing that ACT values were slightly higher than STM results, which is consistent with Zaar/Simonsen et al.^35^. This result demonstrates the equivalence and agreement of both benchmark tests despite the substantial difference in testing time (Figure 3, right panel), with ACT requiring much shorter testing time in line with Zaar/Simonsen et al.^35^

### Efficacy of non-verbal instructions

To evaluate the efficacy of non-verbal instructions, a test run in *block* 1 was considered successful if the ACT value obtained was within 4 dB of the average of results obtained in *block*s 2-4. The decision to use 4 dB as a criterion was taken because it is two steps apart in the clinical procedure (employing a stepsize of 2 dB) and is considered within the limits of agreement of the test^35^. *Block* 1 served as an experimental *block* where some participants were expected to perform less well due to the absence of training or previous explanations and lack of familiarity with the manual ACT test at this point in time.

Table 2 shows the success rate of each U-ACT test candidate across the three participant groups. The NH group demonstrated high success rates for most U-ACT candidates, while the HI sub-groups showed lower performance, particularly those with lower cognitive abilities HI_lo_ (45-63%). Notably, the 3-AFC task (U-ACT-3 and-4) achieved the highest success rates across both HI groups. For those paradigms, 7/11 participants with lower cognitive abilities and 9/11 with higher cognitive abilities performed reliably without prior knowledge and only with the help of the non-verbal instructions. The test with the lowest success rate in the two HI sub-groups was U-ACT-2 while the U-ACT-1, with running stimulus, showed low success rate only in the group with lower cognitive abilities. Note that U-ACT-2 is the only candidate that has catch trials.

**Table 2:** Success rates obtained with the different candidate paradigms in the three participant groups. Definition of success: U-ACT-x in block 1 was less than 4 dB apart from the average U-ACT-x result obtained in the three subsequent blocks.

|  | U-ACT-0 | U-ACT-1 | U-ACT-2 | U-ACT-3 | U-ACT-4 |
| --- | --- | --- | --- | --- | --- |
| NH | 100% | 100% | 100% | 83% | 100% |
| HI<sub>hi</sub> | 72% | 63% | 54% | 81% | 100% |
| HI<sub>lo</sub> | 54% | 45% | 54% | 63% | 63% |

Participants frequently utilized support features during the first *block*. In 31% of the test runs (44/140), participants accessed written instructions; 18% (25/140) required additional visual cues, and 11% (15/140) requested assistance from the examiner. The majority of’call the examiner’ requests were attributed to the user interface (UI) design at the end of the demonstration phase, where some participants repeated the demo multiple times after failing to locate the’start test’ button. By the second visit (Block 3), support requirements decreased: written instructions were accessed in only 13% of runs (18/140) and visual cues in 5% (7/140), with zero instances of examiner intervention.

### Effect of cognitive abilities, task, and procedure on the measured thresholds

To explore the effect of *task*, *procedure*, and the likely influence of cognitive abilities, we analyzed the differences between the thresholds obtained from all five U-ACT test candidates and those from the manual ACT, with which the participants were also tested. Figure 4 shows the threshold differences for each of the three groups of participants. The 1-button tasks (U-ACT-0 and-1) with running stimuli showed minimal differences compared to the manual ACT values. In contrast, tasks involving a 3-interval sequence (U-ACT-2-4) yielded significantly lower thresholds than those from the manual test, in agreement with Zaar/Simonsen et al.^35^. In the analysis of variance (ANOVA) with a mixed effects model, no significant differences were observed across the participant *groups* suggesting that any differences in mean performance among ACT and the various U-ACT tests were largely independent of the participants’ cognitive abilities.

**Figure 4:**
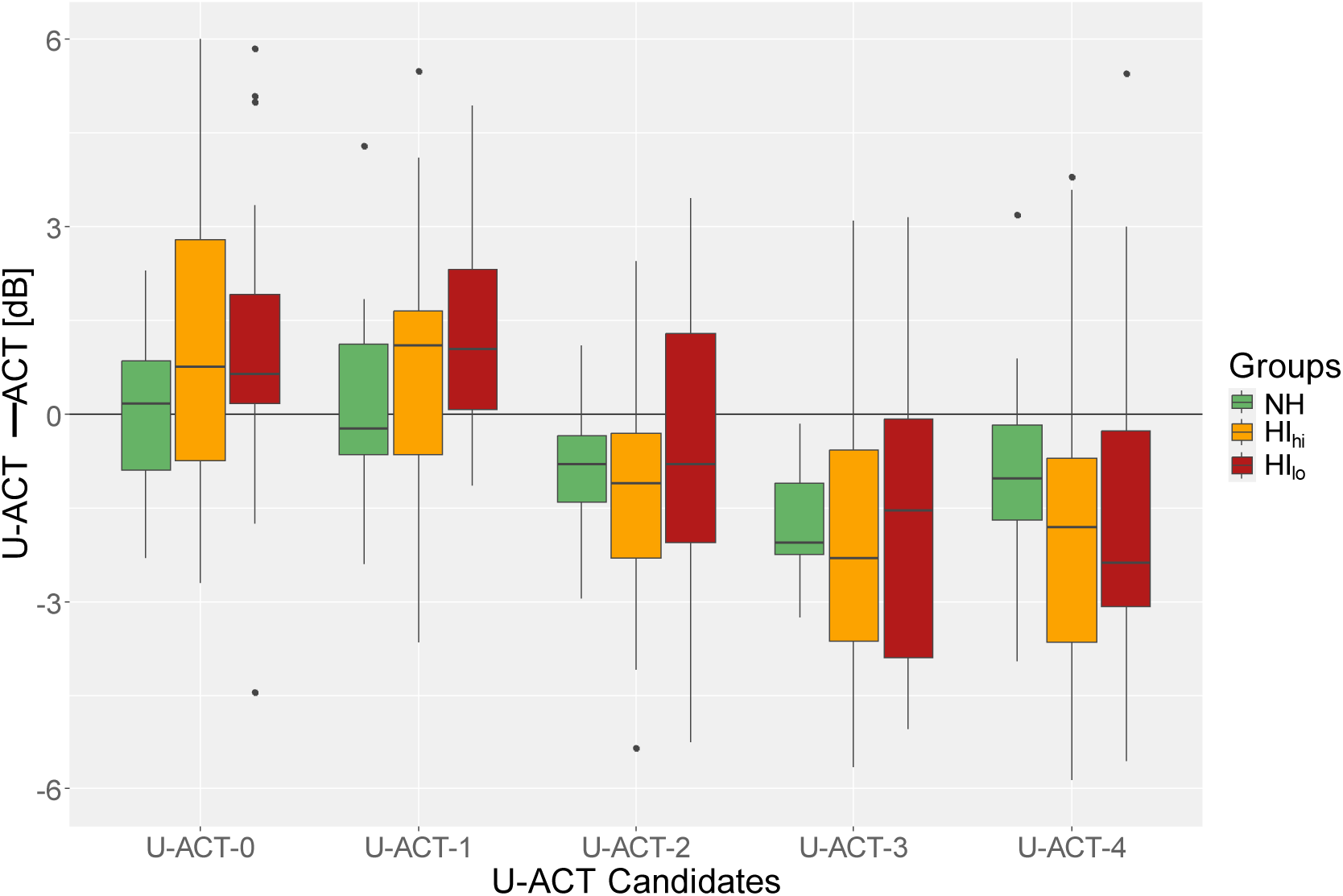
Differences between user-operated Audible Contrast Thresholds (U-ACT) and manually obtained Audible Contrast Thresholds (ACT). In the boxplots, the horizontal lines indicate the median, boxes indicate interquartile difference, and vertical lines indicate the range between the max and minimum values within ±1.5 times the interquartile range (whiskers). Individual data points outside of the whiskers are also indicated.

Examining the overall data*, task*-specific differences were evident: there were significant mean threshold differences between the 1-button tasks (U-ACT-0 and U-ACT-1) and the 3-interval yes/no choice of U-ACT-2 (t = 8.96, p < 0.001) and between the 1-button tasks and the 3-AFC tasks, i.e., U-ACT-3 and-4, (t = 15.17, p < 0.001). No significant differences were observed among the tests with sequential presentation (U-ACT-2, U-ACT-3 and U-ACT-4). Notably, the self-paced sequential presentation allows the participant to reflect on the response after the presentation of the three intervals, while the 1-button task is done while there is a running stimulus and it has a fixed response window (0.2-1.6 seconds after the onset of the target stimulus).

### Quality of measurements: Reliability and agreement with benchmarks

The test-retest reliability of the U-ACT test candidates was evaluated using the thresholds obtained in the second *block* of each visit (that is *block* 2 in visit 1 and *block* 4 in visit 2).

For the analysis of the reliability and repeatability, three different metrics were employed: the inter-class correlation (ICC)^44^, the coefficient of repeatability (CoR)^45^ from the Bland-Altman plots^46,47^ and a metric based on information retrieval named Fractional Rank Precision (FRP)^48^. While ICC and FRP are quantified on a scale from 0 to 1, where 1 indicates ideal reliability, the CoR represents the range within which repeated measurements are likely to fall. The CoR is thus expressed in dB, and a smaller range indicates a more reproducible measure. These metrics provided a comprehensive assessment of the tests’ consistency.

Table 3 shows the reliability results of the U-ACT test candidates and the two benchmarks. The STM test demonstrated the highest reliability. U-ACT-2 emerged as the second-best test, and thus the best U-ACT candidate in terms of reliability, across all considered metrics. The manual 1-button ACT was substantially more reliable than its user-operated counterparts (U-ACT-0 and U-ACT-1).

**Table 3:** Test-retest reliability of the user-operated Audible Contrast Threshold (U-ACT) test candidates and test benchmarks analyzed using data from blocks 2 and 4. The three metrics are the inter-class correlation (ICC), coefficient of repeatability (CoR), and fractional rank precision (FRP). The rightmost column summarizes the average test duration. Bold letters indicate the best values among the U-ACT test candidates.

| *Test paradigm* | ICC (2,1) | CoR<br>(dB nCL) | FRP<br>(ratio 0-1) | Average Duration<br>(s) |
| --- | --- | --- | --- | --- |
| U-ACT-0 | 0.71 | 3.72 | 0.75 | **90** |
| U-ACT-1 | 0.69 | 4.11 | 0.70 | 97 |
| U-ACT-2 | **0.84** | **3.27** | **0.78** | *202 |
| U-ACT-3 | 0.80 | 3.59 | 0.73 | *145 |
| U-ACT-4 | 0.78 | 3.62 | 0.74 | *292 |
| ACT | 0.83 | 3.51 | 0.75 | 82 |
| STM | 0.87 | 3.04 | 0.75 | 324 |
| * In hindsight, U-ACT-2,3 and 4 included an unnecessary 1-second pause, which added about 25 seconds to the reported test duration. |  |  |  |  |

To assess the agreement between U-ACT candidates and benchmarks, the same metrics were applied to the average thresholds obtained across *blocks* 2, 3, and 4. Table 4 highlights that the U-ACT-2 test candidate showed remarkable agreement with both benchmarks ACT and STM achieving an ICC > 0.9, a low coefficient of repeatability (lower than the one corresponding to ACT, i.e., CoR < 3.5) and the best fractional rank precision (FRP = 0.8) compared to the other candidates. Although the other U-ACT test candidates showed an acceptable agreement with both benchmarks, it is remarkable that U-ACT-4 exhibited an excellent agreement with the STM test (ICC(2,k) = 0.97) despite being shorter and using larger step sizes (see Methods).

**Table 4:** Agreement between the U-ACT test candidates and the test benchmarks analyzed using the mean across blocks 2, 3, and 4. The three metrics were the inter-class correlation (ICC), coefficient of repeatability (CoR) and fractional rank precision (FRP).The best value for each metric is highlighted in bold font.

|  | ICC (2,k) |  | CoR<br>(dB nCL) |  | FRP<br>(ratio 0-1) |  |
| --- | --- | --- | --- | --- | --- | --- |
| Agreement | ACT | STM | ACT | STM | ACT | STM |
| U-ACT-0 | 0.86 | 0.69 | 3.46 | 4.18 | 0.77 | 0.69 |
| U-ACT-1 | 0.88 | 0.76 | 3.70 | 4.37 | **0.78** | 0.66 |
| U-ACT-2 | **0.93** | 0.94 | **3.16** | 2.90 | 0.77 | 0.82 |
| U-ACT-3 | 0.80 | 0.95 | 4.15 | 2.81 | 0.66 | 0.81 |
| U-ACT-4 | 0.86 | **0.97** | 3.77 | **2.43** | 0.69 | **0.83** |

### Quality of interaction

Figure 5 and Figure 6 display the standardized data (see Methods) of the usability evaluation for the five U-ACT candidate tests.

**Figure 5:**
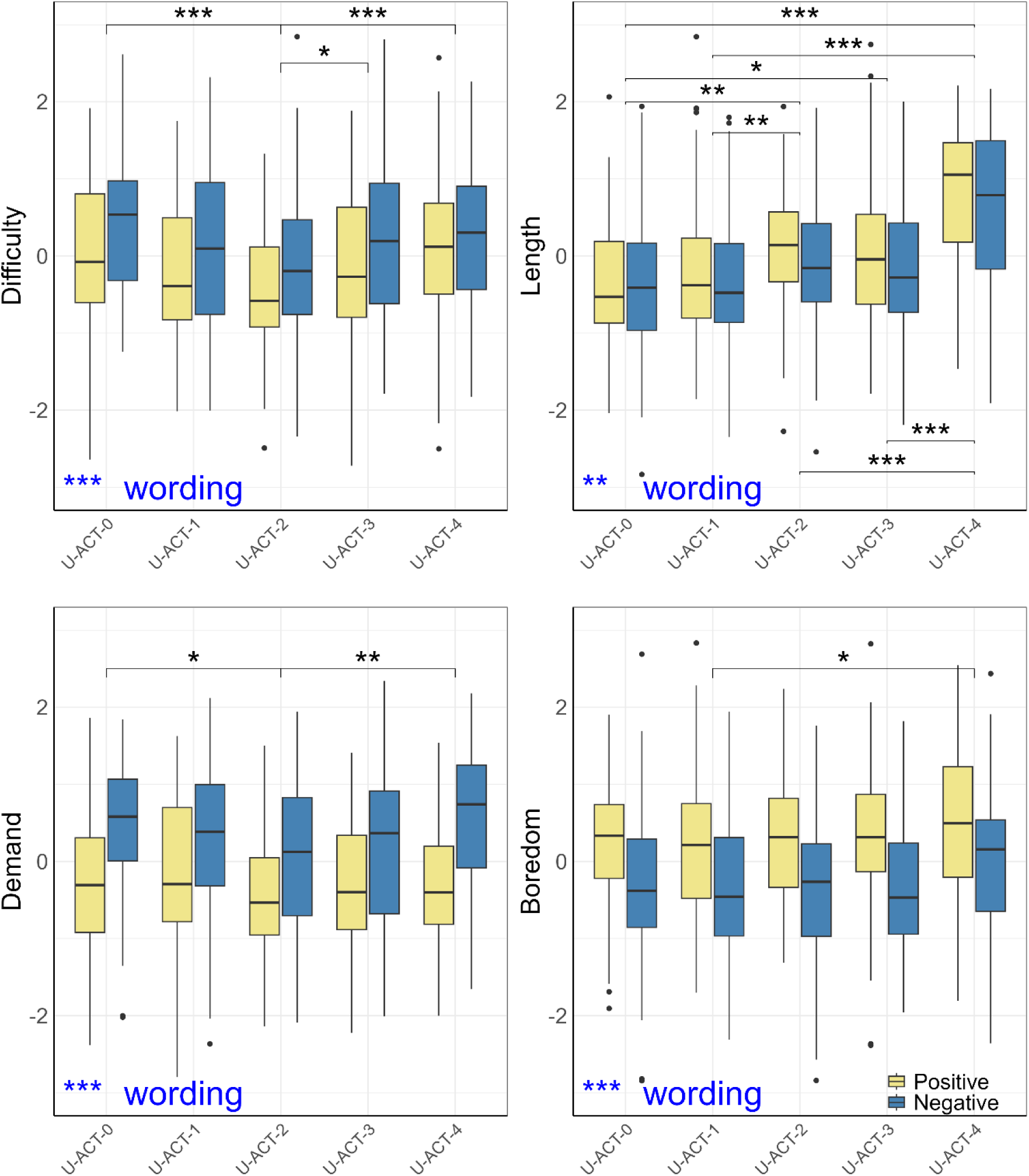
Subjective evaluation of the usability of the user-operated Audible Contrast Threshold (U-ACT) candidate tests (U-ACT-0 to U-ACT-4). Standardized subjective ratings of the attributes related to the test: Difficulty, Length, Demand, and Boredom. Boxplots depict the median, interquartile ranges, and whiskers as in Figure 4. The colors represent Positive (yellow) or Negative (blue) wording. Asterisks indicate statistically significant differences between groups (* p < 0.05, ** p < 0.01, *** p < 0.001). The significance of the attribute wording is shown at the bottom left corners.

**Figure 6:**
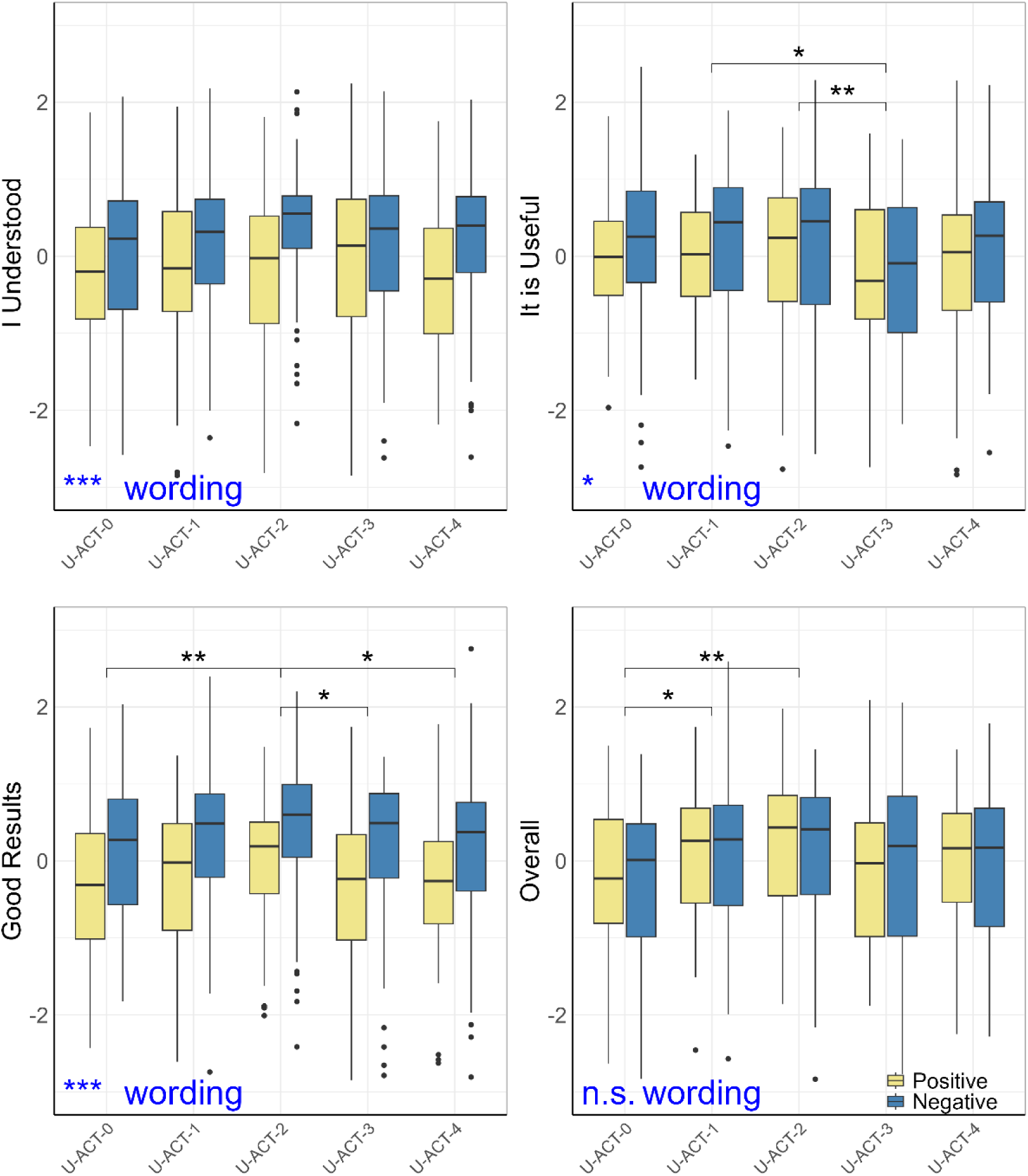
Evaluation of the usability of the user-operated Audible Contrast Threshold (U-ACT) candidate tests. Average subjective ratings of the attributes related to self-confidence with the tests: Understanding of Task, Usefulness, Goodness of Results, and Overall opinion. Boxplots depict the median, interquartile ranges, and whiskers as in Figure 4. The colors represent Positive (yellow) or Negative (blue) wording. Asterisks indicate statistically significant differences between groups (n.s. p>0.05, * p < 0.05, ** p < 0.01, *** p < 0.001).

Figure 5 compares subjective ratings of attributes related to the test experience. Lower ratings indicate better results, meaning less difficult, shorter, less demanding, or less boring. The *wording*, which reflects a positive or negative formulation of the attribute, had a significant impact on all four attributes. This suggests that the phrasing of statements can affect ratings. For example, participants found U-ACT-2 to be less “difficult” than U-ACT-4 but still equally “easy”. The most significant results showed that U-ACT-4 was perceived as significantly longer than the other tests and U-ACT-2 was less demanding than U-ACT-0 and U-ACT-4.

Figure 6 shows subjective ratings reflecting attributes related to participants’ confidence with the tests. Higher ratings indicate better results, meaning that the participant understood the task, found the test useful, believed they did well, or had a positive overall opinion of the test. From these attributes, only the overall opinion did not differ significantly in terms of *wording*. U-ACT-3 was perceived as significantly less useful than U-ACT-1 and U-ACT-2. Additionally, participants reported that the results obtained using U-ACT-2 were significantly better than the ones from U-ACT-0, U-ACT-3 or U-ACT-0.

## Discussion

The present study consisted of a multi-dimensional evaluation of various test candidates for a user-operated auditory test. The study aimed to generate quantitative data on test reliability, observations from tracking user interactions with the system, and users’ self-reported opinions about usability. Instructions are critical for obtaining meaningful results in user-operated tests. Initial results from *block* 1, conducted without additional instructions, indicated that a substantial proportion of hearing-impaired listeners may struggle with understanding their task, particularly those with lower cognitive abilities. All test candidates showed good agreement with benchmarks and good test-retest reliability. While the procedures and tasks differed from classical psychoacoustic tests^28,29^, results were comparable to benchmarks in terms of perceptual evaluation results. Some insights emerged, such as perceived differences in test difficulty and length among candidates, and a difference in results based on attribute wording, which showed a strong and clear significance in user evaluations.

### The chosen test candidate

The study evaluated user interaction in a realistic scenario and aimed to identify the “best candidate” for user-operated ACT. The selected candidate was U-ACT-2, which exhibited high reliability and repeatability, with excellent agreement compared to ACT and STM benchmarks. Overall, the test was appropriate in terms of length and acceptable, in the sense that the users’ evaluation of U-ACT-2 was average or better than average in all dimensions considered. While generally easy to perform due to its simplified target/reference discrimination task, U-ACT-2 initially presented comprehension challenges for some participants during the first block. However, addressing this through improved instructions and combined verbal/non-verbal cues might be sufficient^49^.

### Relation to other studies

Previous studies have proposed quick and reliable automated tests of spectro-temporal modulation perception^50–52^. In most cases, the process of developing and validating the test has been meticulously done in various steps, adding complexity in each new study towards a realistic application. For example, a spectral ripple discrimination test was developed and further optimized for clinical applications^50,53^. However, there is no clinical evidence for the use of the test for aural rehabilitation. Evaluations of the STM task in the Portable Automated Rapid Testing (P.A.R.T)^54^ platform by Gallun, Lelo de Larrea-Mancera and colleagues initially focused on stimulus generation^55^ and population-specific performance^56,57^. Subsequent research has significantly expanded this scope to include formal psychoacoustic modeling via the Adaptive Scan algorithm to improve time efficiency and rigorous usability testing to ensure the platform’s viability for at-home auditory assessment^57,58^. In contrast, we have directly tested U-ACT in a realistic environment, with a diverse sample in terms of auditory and cognitive abilities.

The evaluation methods of user-operated audiological tests are mainly focused on accuracy and reliability^59^. The automated pure-tone audiometry protocol utilized in the UAud project (AMTAS) is supported by a substantial body of evidence validating its reliability and clinical effectiveness^36^. Previous multi-center studies have demonstrated high correlation between AMTAS-derived thresholds and traditional manual audiometry^60^, including validations of at-home settings^61^. Furthermore, the UAud project has examined the impact of AMTAS results on hearing-aid fitting outcomes ^62^. However, comprehensive usability data remain scarce, particularly regarding user experience of older adults. This gap is critical, as cognitive decline may significantly and negatively impact the successful completion of user-operated diagnostics in the overwhelmingly elderly target population. The present study addresses this by including older adults with lower cognitive abilities to determine if U-ACT is accessible to a broad population regardless of cognitive profile, evaluating also the use of instructions and the quality of interaction for five alternative test variants. To our knowledge, this is the first study in audiology to select and evaluate an auditory test using a multimodal approach centered on the user experience and guided by the RAMES framework.

### Subjective evaluation

Subjective evaluations are known to have response biases and systematic errors that can distort survey outcomes. In our study, we chose a visual analogue scale because it offers higher sensitivity to small changes than discrete scales and provides continuous data suitable for more powerful statistical analysis^63^. However, it is known to have leniency bias, end-aversion bias, and directional bias (such as pseudo-neglect)^64^. The challenge is to control how the participants make use of the scale to ensure they accurately translate their sensory experiences into the linear format without clustering responses or reinterpreting anchors.

In our study, the inclusion of both positive and negative item wordings was a deliberate methodological choice designed to mitigate acquiescence bias (the tendency of respondents to agree with statements regardless of their content) and directional response bias. By employing dual formulations for the same attribute, we sought to enhance the internal validity of the subjective ratings. A significant difference between U-ACT candidates across both wordings thus indicates a robust perceived difference in usability. In contrast, when the *wording* factor itself is significant, it highlights a framing effect, suggesting that participant judgments are sensitive to how the question is posed.

Furthermore, by doubling the observations per attribute through these dual formulations, we increased the statistical power of the usability evaluation.

The response bias has been investigated in several studies and different mitigation approaches have been tried, such as using intermediate ticks in the scale^65^, combination with other techniques^64^, or complex statistical modelling^66^. In our study, the combination of standardization of the data and the redundancy obtained through use of different wording allowed us to identify subtle differences between the test candidates.

### Strengths and limitations of the study

The UAud approach aims to implement user-operated hearing assessments in healthcare services. We proposed a protocol for multi-dimensional evaluation involving reliability, efficiency, efficacy, and subjective evaluations. Key strengths of the study include a diverse participant selection regarding hearing losses and cognitive abilities, and a comprehensive evaluation using a simple two-visit protocol.

While the multi-dimensional evaluation was valuable, there are limitations to consider. Firstly, participants were sourced from the university’s database, potentially biasing results due to prior hearing research experience affecting task interactions. Besides, the cognitive screening was only based on one aspect of a specific ability (i.e., working memory).

Although other types of assessments were considered^67,68^, we decided to use a robust test that has been previously used at the Technical University of Denmark (DTU) in various studies^40^, rather than a test including aspects like attention or executive function that could be time consuming or less reliable. Secondly, repeated assessments of the same listening ability within each block may have positively influenced results through training effects.

This was a known limitation that we could not disentangle in the analysis. As an alternative, we could have used a different experimental design such as a randomized trial with one group per test candidate, however, this approach would have resulted in other limitations since each individual would only have used one of the candidates. Additionally, the protocol lacked speech intelligibility in noise tests as a reference measure, as included in Zaar et al.^34^. The U-ACT test is expected to be equivalent to the clinical ACT and have a similar predictive power so it can be considered a proxy for speech-in-noise tests.

However, this should be explored in a separate study comparing the two contrast tests with the hearing-in-noise test with a clinically relevant population.

### Outlook

The auditory tests used for audiological diagnosis should provide a reliable estimate of a patient’s hearing status; however, manual and user-operated tests need not be based on identical tasks or procedures^6^. While the reliability of test results is influenced by user behavior, it is the responsibility of both the audiologist (human) and the system (represented by the system designer) to present the test in a manner that minimizes influencing factors unrelated to hearing status, such as attention, understanding of the task, and fatigue^69,70^. In user-operated tests, it is reasonable to think that this difference in the allowed response time is the most influential factor. The results obtained here support the use of sequential presentation in user-operated tests (as also demonstrated in AMTAS studies^36^), while emphasizing the importance of the examiner’s role in manually operated ACT. In research, the current version of U-ACT opens the possibility to test other populations (pediatric, cognitively impaired) using a single and similar framework for audibility and audible contrast tests, and to explore the use of U-ACT as a remote test (as in the case of the audiometry^71^). Overall, U-ACT can help to further investigate the role of contrast loss in auditory perception and communication handicap.

## Methods

### Study design and participants

#### Participant recruitment

The potential participants of the study were recruited via purposive convenience sampling from the database of the Hearing Systems Section at DTU. In the recruitment process, care was taken to ensure a balanced number of male and female participants, and a reasonable variety of hearing losses and cognitive abilities as reflected in their audiometry and RDS scores. The potential participants were divided into three groups already during the recruitment based on the information contained in the database. The study aimed to include a total of 30 participants, stratified into three groups: six young NH controls, twelve HI listeners with high cognitive abilities (HI_hi_), and twelve HI listeners with low cognitive abilities (HI_lo_). We intentionally recruited 6 NH listeners below 30 years old which is sufficient to establish a stable baseline for interaction quality ^72^. Cognitive classification was based on the RDS scores; the high-ability group included scores from 14 to 21, while the low-ability group included scores from 6 to 12. The only exclusion criteria were a hearing loss above the range where the ACT can be performed (i.e., exceeding 95 dB HL) or an inability to operate a tablet due to dexterity or vision problems. Thus, a total of 30 participants with diverse hearing and cognitive abilities were recruited for the study. Two participants abandoned the study after the first visit and 28 participants completed the protocol. This study was carried out in accordance with the ethical approval of the Danish Science-Ethics Committee granted by the Capital Region Committee (H-16036391). All participants gave written informed consent and were offered an economic compensation for their participation.

### Equipment and testing environment

The study was conducted in the laboratory of the Hearing Systems Section at the DTU, specifically in the audiology clinic (which resembles a typical audiology clinic). This facility includes a sound-isolated booth equipped with a clinical audiometer, videoscope, middle-ear analyzer, and computer for conducting hearing-in-noise tests and cognitive assessments. For the present study, we employed a clinical research prototype of Audible Contrast Threshold testing implemented in MATLAB^®^ (Version 9.8, The MathWorks Inc.). The prototype consists of an RME UC soundcard (RME Audio, Haimhausen, Germany) connected to a Lake People G103-P MKII amplifier (Lake People, Überlingen, Germany). The prototype included a pair of headphones Radioear DD450 (Middelfart, Denmark), and a push button. For the user-operated tests, we used an iPad 8^th^ generation MYL92KN/A (Apple Inc., Cupertino, CA, USA) connected to the computer functioning as an external screen using the software Splashtop Wired Xdisplay (Splashtop Inc., San Jose, CA, USA).

### Protocol

The participants were invited for two visits that occurred at least one week and no more than four weeks apart. In the first visit, they were introduced to the study and were informed that the test involved a usability test and that we wanted the participants to interact with the system. After this briefing, participants underwent a standard pure-tone audiometry examination administered by a trained audiologist before they conducted the main experiment. The second visit began with the administration of the RDS test followed by the main experiment. The main experiments consisted of a total of four testing *block*s (two *block*s in each visit), where participants evaluated five alternative U-ACT candidates, with candidates presented in balanced order across groups and *block*s. In each visit, both test benchmarks (that is, ACT and STM) were administered in between the two testing *block*s once the examiner had explained the test to the participant. After *block* 4, semi-structured interviews were conducted with each participant. The interview included questions about the opinions of having user-operated tests in the clinic and the utility of the U-ACT to complement the audiometry. The qualitative results of these interviews are not within the scope of the present investigation.

### Auditory tests of audible contrast sensitivity

#### Stimuli

The stimuli used in all the tests performed in the study were identical to those used in the ACT as described by Zaar/Simonsen et al.^35^, which has been thoroughly explained in previous studies using STM detection^34^. In general terms, the stimuli consist of 1-second-long broadband noise carriers (354-2000 Hz) – serving as reference stimuli-, onto which spectral modulations of 2 cycles per octave and temporal modulations of 4 Hz are imposed to create the target stimuli. The experimental variable was the modulation depth (m), which was transformed into dB FS by 20 𝑙𝑜𝑔(𝑚) for the STM test, and into dB nCL for all the ACT tests (both manual and user operated).

#### Test benchmarks

Clinical assessment of contrast sensitivity has emerged as an important diagnostic tool for evaluating a patient’s ability to extract subtle acoustic information from complex sounds, and its use in hearing rehabilitation is supported by evidence^34,35^. In this study, we compare the performance of U-ACT candidates with two benchmarks that have been previously used in relevant studies^34,35^. The first benchmark, referred to as the STM detection test^28,34^, is a psychoacoustic measure that utilizes a three-alternative forced choice (3-AFC) task and a 2-down 1-up procedure that approximates the 70.7%-point on the psychometric function^29^. The psychometric function represents the probability of detecting the target sound and it is often modelled as a logistic regression function with a midpoint (threshold) and a slope that define the performance for any level of the tracking variable. The second benchmark is the clinical version of the Audible Contrast Threshold (ACT™)^35^ test, which has been widely used in previous studies.

#### Test tasks

Three *task*s were considered for the U-ACT test candidates. These were:

1. 1-button task: The stimuli are presented in 1-second-long successive waves of noise, and occasionally the wave contains the target stimulus (i.e. it is modulated), thus differing from the other (unmodulated) reference waves. We use the term ‘wave’ because the 1-second-long noise has a fade-in and fade-out and it is followed immediately by another 1-second-long noise so it sounds like waves in the ocean. A large virtual button, referred to as the’1-button’, was placed on the touch screen occupying 40% of the screen’s area at the bottom. We implemented a fixed response window (starting 200 ms after stimulus onset and ending 600 ms after offset, for a total of 1.6 s) to categorize trials as HIT or MISS. This window mirrors the parameters used in the manual ACT^35^, assuming comparable response times between physical buttons and touch screens. If the participant did not touch the screen, the trial was marked as MISS, whereas the trial was marked as a HIT if the participant touched the screen during the response window. The screen touches outside of the response window were registered as false positives. This task was used in U-ACT-0 and U-ACT-1.
2. Yes/No task: The stimuli were presented in triplets, the first and last intervals were always references, while the second interval could be either a reference or a target stimulus. The user’s task was to press either the button depicting an ambulance when detecting a target, or the button depicting a wave indicating the detection of a reference, i.e., no detection of a target. This task resembles the one used in AMTAS^36^ and includes catch trials with feedback when the participant gets caught pressing the button when there was no target stimulus presented. This was done to ensure that the performance and attention is maintained during the test. The catch trials consisted of trials containing a reference stimulus in the second interval. There were 5 catch trials which were assigned at the beginning of the test to be presented before specific trials. The first catch trial is always between the first and second trials, the remaining were assigned randomly. The feedback was presented only when the participant was caught in a catch trial which is expected to minimize the bias and have less influence on the results of the actual test ^73,74^. There were no restrictions on response time since the stimuli were presented sequentially. This task was used in U-ACT-2.
3. 3-AFC task: The stimuli were presented in triplets. The screen showed three virtual buttons with numbers 1-3, which were highlighted sequentially in synchrony with the playback of the three intervals. The target stimulus was randomly placed in one of the three intervals. The task of the participant was to indicate which of the three intervals was different from the other two by pressing the button numbered with that interval. If no differences were heard, guessing was compulsory to advance the test. There were no restrictions on response time since the stimuli were presented sequentially. This task was used in U-ACT-3 and U-ACT-4.

### Test procedures

Three *procedure*s were considered for the U-ACT test candidates:

- Modified Hughson-Westlake procedure^42^: This procedure was identical to that used in the ACT test, but automatic. After each response (either HIT or MISS), the contrast level of the next target stimulus was adjusted by, respectively, decreasing or increasing the contrast level. The step size was 2 dB nCL. After a HIT, the contrast level was decreased by 2 steps (i.e., 4 dB nCL). After a MISS, the contrast level was increased by 1 step (i.e., 2 dB nCL). Following the’ascending method’^75^ used in pure-tone audiometry, testing ceased when three out of five positive responses occurred at a single contrast level, provided each followed a non-response. This procedure was used in U-ACT-0.
- Custom Bayesian procedure: The presented procedure is an adaptation of the BAYES-FIG^76^. In their procedure, Remus and Collins combined Bayesian inference with the Fisher information matrix and Theory of Optimal Experiments. Overall, the Bayesian inference aims to reduce the uncertainty by obtaining new evidence (a response at a certain level) and updating the probability of a given combination of midpoint and slope of the psychometric function to be the true result. The Fisher information matrix is obtained using the derivative of the psychometric function with respect to the midpoint and the slope and estimating which value of the tracking variable is the one that should be tested next to maximize the information provided by the new evidence. However, our method incorporates distinct variations:

`o` Bayesian updates: Unlike BAYES-FIG, our method utilizes the last two trials (n-1 and n) to update probabilities, rather than relying solely on the last trial as in other psychoacoustic procedures based on Bayes theorem. This was done under the assumption that using two trials makes the estimation more robust to momentary lapses in participant attention. This assumption was tested and confirmed using Monte Carlo simulations.
`o` Informational gain function: Instead of using Fisher’s informational gain, our method employs a different information gain function that is derived solely from the slope’s derivative.
`o` Optimal next CL algorithm: In the BAYES-FIG the selection of the level for the next trial is based on the maximum of the information gain function. In contrast, our method alternates between the maximum and minimum values corresponding to the 82% and 18% points of the psychometric function, respectively, on even and odd trials. The exact points of the psychometric function are defined by the partial derivative of the psychometric function with respect to the slope.
`o` Exception inclusion: To mirror the manual test experiences reported by audiologists, our procedure incorporates additional catch trials (used in the Yes/No task) or presentations above threshold to maintain patient attention and ensure consistent performance. These presentations occurred based on erratic patient responses that might suggest that the patient has lost the internal reference of the sound that they have to detect.
- Catch Trials: In the Yes/No task, catch trials were included in the procedure. By default, the system considers 4-5 catch trials assigned randomly to specific trial numbers with 4-8 trials in between.

Overall, the Bayesian procedure consisted of 25 trials that started with 8 pre-test trials for a first approximation of the threshold using the modified Hughson-Westlake method and then continued alternating easier and more difficult presentations using the Optimal next CL algorithm. This procedure is employed in U-ACT-1,2 and 3.

- Modified 2-down 1-up procedure: This procedure was based on the 2-down 1-up described by Levitt^29^, with a slight modification to make it shorter. Thus, the contrast level of the stimulus was decreased only if there were two consecutive correct responses and increased otherwise. The entire procedure consisted of two iterations (sub-runs), each of them consisting of 4 upper reversals with decreasing step size. An upper reversal is a change of direction that takes place in ascending direction after two consecutive positive responses. At the start, the step size was 8 dB after the first upper reversal it decreased to 4 dB, and after the second to 2 dB. The procedure then continued until there was a correct response after a negative response. The second iteration started 4 dB nCL above the last presentation level and the procedure was repeated identically. This procedure was employed in U-ACT-4

#### Post-processing and threshold estimates

For each test run, the levels of the tracking variable and the participant responses constituted a trace. In the context of adaptive testing procedures, a trace is a step-by-step history of the tracking variable from the start and until it converges to the threshold. The trace was fitted to a psychometric function^77^:

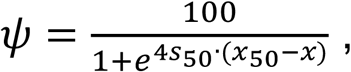

where the slope (𝑠_50_) and midpoint (𝑥_50_) are the parameters, 𝑥 is the contrast level and 𝜓 is the % correct. In the fitting process, the slope was kept fixed (𝑠_50_ = 0.2518 dB^−1^) and 𝑥 was defined in the closed interval [-8, 16] with a step size of 0.1. The variable 𝑥 was thus allowed to go below the minimum value used with ACT (-4 dB nCL) to ensure accurate representation of the entire psychometric function, particularly for cases where threshold levels were very low. For all the ACT tests (manual and user-operated) the ACT value was taken as the 𝜓 = 70.216 %-point on the psychometric function, following the recommendations from Zaar/Simonsen et al.^35^. The STM benchmark followed the same procedure as in the Zaar et al.^28,34^ studies (i.e. a transformed 2-down 1-up procedure^29^ with a final step size of 1 dB). However, the test procedure of the STM was extended by 8 more reversals compared to the studies of Zaar et al.^28,34^ which allows an investigation of the likely effects of fatigue (not considered here). The threshold estimate (𝑆𝑇𝑀_𝑡ℎ_) was the resulting mean across the values of the first 8 reversals (4 upper and 4 lower) at a step size of 1 dB In this case, the threshold corresponds to the 𝜓 = 70.7%-point^29^.

### Non-verbal instructions

The instructions consisted in a succession of assignments and were designed so they only contained non-verbal audiovisual information^78^. The participants had to go through four instruction phases, each of which comprised a specific assignment and a goal that they had to achieve by themselves. However, the participants could make use of different types of support during the instruction phases. 1) Help: If the user pressed a question mark in the bottom left corner of the touch screen, minimalistic written information was added to the screen. 2) Written instructions: If the user pressed the blue “information” button, thorough written instruction appeared on the screen. 3) Call the examiner: The user always had the option to call the examiner for task explanation. The participants could go backwards and forwards and were not obliged to complete the assignment that was presented in each step of the instruction phase. A cartoon of the instructions and main visual elements is depicted in Figure 7.

**Figure 7:**
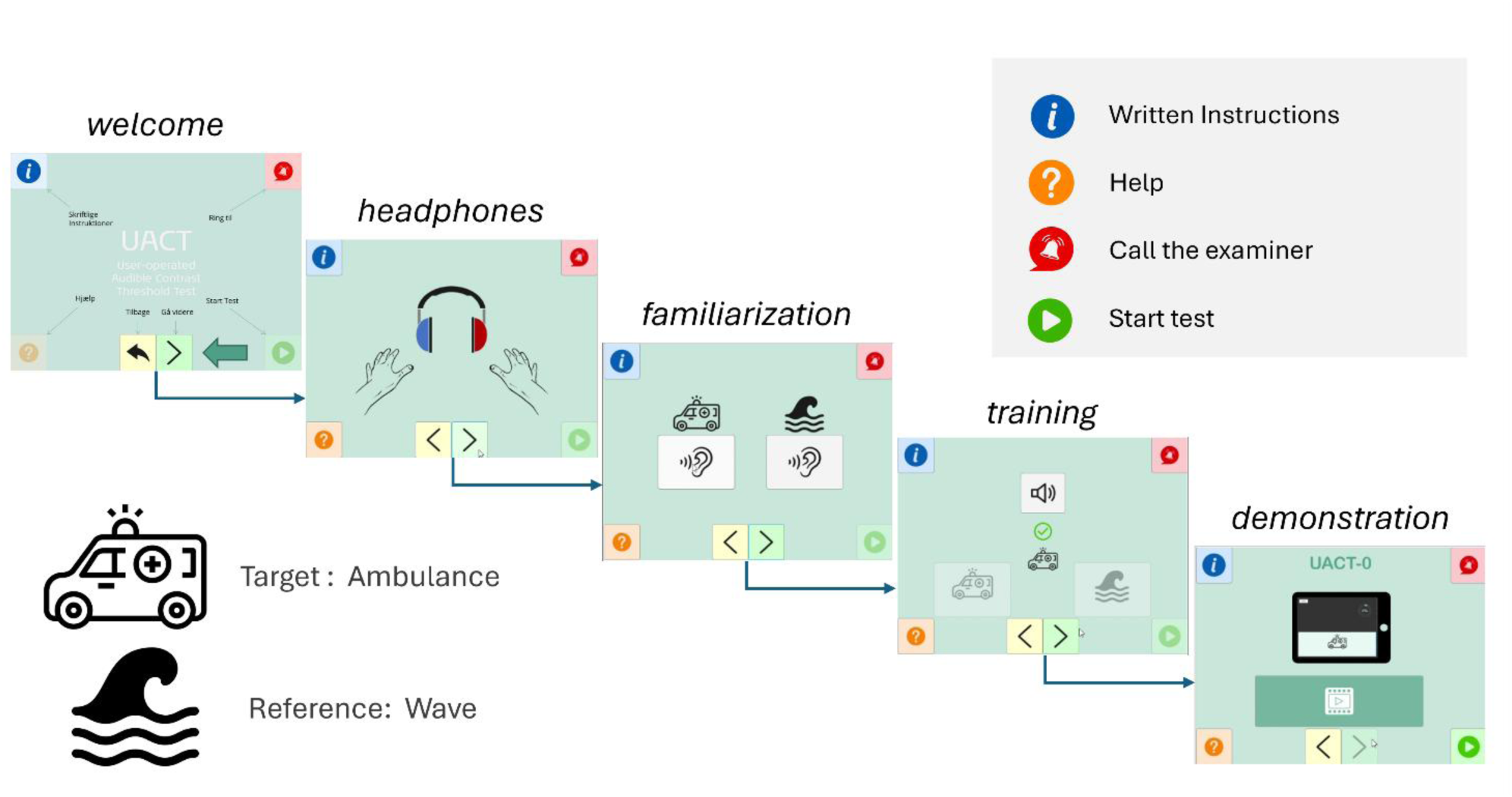
User workflow for the U-ACT test. The participant progresses from a welcome screen through headphone setup, sound familiarization, and training, before starting the test demonstration. The interface uses an “Ambulance” as the target-sound icon and a “Wave” as the reference icon, with on-screen help always available. The participant can make use of 3 types of help: extra information, written instructions, or call the examiner to obtain verbal instructions.

The instructions aimed at achieving four goals for the participant:

1. Correct headphone placement. The screen displayed an image of the headphones indicating which side corresponds to which ear.
2. Become familiar with the stimuli: The screen showed an ambulance and a wave, each of them with a button with an icon representing a loudspeaker.
3. Try out a detection test. The screen showed three buttons. The button with a speaker was enabled and played a sound when pressed. The participant was then required to press one of the other two buttons (ambulance or wave). After the button was pressed by the participant, a sign appeared on the screen indicating whether their choice was correct or incorrect.
4. Watch a demonstration. The participants should watch a short video showing how to perform the *task*.

The use of the instructions was investigated in *block* 1. All the events (i.e., next phase starts, go forward, go back, etc.), button presses, and response times were captured by the system.

### Subjective evaluation

The evaluation was carried out using the touch screen. The data collection process involved the use of a visual analogue continuous scale (VAS), following each run in *block*s 2, 3, and 4. On the screen, a statement was written starting always with “The test was…” and below it, a continuous slider served to collect the judgments. To mimic a Likert scale, the slide had the three labels: “strongly disagree”, “neutral” and “strongly agree” and the result was a number between-50 and 50. The collected attributes included difficulty, self-perceived duration, demanding, boredom, as well as evaluations on the test’s outcome such as understanding the instructions, usefulness, goodness of results, and overall impression. These attributes were assessed twice: once using positive statements like “The test was easy”, and another time employing negative statements such as “The test was difficult”.

### Analyses

#### Statistical analysis of threshold differences

All statistical analyses were carried out using linear mixed-effects models and based on an analysis of variance (ANOVA) with subsequent post-hoc analyses of pairwise differences. The analyses were run in RStudio (Version 12; Posit team, 2024) using the packages *`lmerTest`* and *`emmeans`*. More details about the investigations can be found in the supplementary material.

For the investigation of the training effects and the effects of *task* and *procedure* on the thresholds, we made use of the difference between the U-ACT test candidate results and the benchmark ACT value of each individual participant (denoted ΔACT). This allowed a fairer comparison of the training effects. For this purpose, *block* 1 was disregarded since the purpose of that *block* was to evaluate the instructions and the participants did not have any previous experience with the test candidates at this point. The effect of the remaining repetitions was included in the factor *block*.

Statistical models included all the possible fixed effects and their interaction with the participant `group` while participant `ID` was included as a random effect. The models had the following form in R notation:

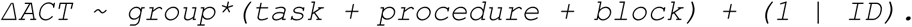

Estimated marginal means (EMMs) for the fixed factors were calculated with Satterthwaite’s degrees-of-freedom approximation. Pairwise contrasts between task conditions were evaluated using Bonferroni-corrected t-tests to account for multiple comparisons. In case of significant interactions, an analysis of the subset of the data related to specific conditions or groups was performed.

#### Test-retest reliability metrics

To evaluate the test-retest performance of the test candidates and the agreement with the test benchmarks, the following metrics were considered:

- Intra-Class Correlation (ICC)^44,79^: This metric is commonly used in test-retest reliability studies. It measures consistency and agreement across different occasions, conditions or procedures, on the same subjects and it makes use of mixed models. For the measure of absolute test-retest reliability, we employed a 2-way random effect model (2 different measurements) with a single rater measurement (the measurement is not the average of various repetitions). In the manuscript we refer to this as ICC(2,1). For the comparisons between tests, we employed a 2-way random effects model, to quantify the absolute agreement, among multiple repetitions. In the manuscript we refer to this as ICC(2,k)^44,79^.
- Mean difference and Coefficient of Repeatability (CoR)^45,47^: These are obtained from Bland & Altman plots used, for example, in the middle panel in Figure 3, where the mean difference represents the bias, and the CoR defines the limits within which 95% of the differences between paired measurements lie. Mathematically, 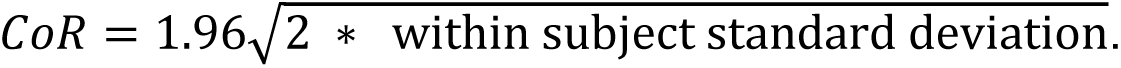
- Fractional Rank Precision (FRP)^80^: This is a metric used to assess the precision of a method in ranking objects or samples and thus related to the mean average precision, which is a standard tool for information retrieval. The higher the FRP value, the better the precision in rankings.

#### Postprocessing and data modelling of subjective evaluation data

Prior to the analysis, data from the subjective evaluations underwent preprocessing to account for variations in the use of the VAS scale among participants and to focus on the differences between U-ACT test candidates. To standardize the ratings, each raw judgement (𝑟𝑎𝑡𝑖𝑛𝑔_𝑖,𝑏_) was transformed using a participant-*block*-specific Z-score:

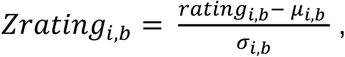

where 𝜇_𝑖,𝑏_ and 𝜎_𝑖,𝑏_ represent the mean and standard deviation, respectively, calculated across all rated attributes but separately for each participant 𝑖 within each experimental *block* 𝑏..

Additionally, both types of wording for each attribute (positive and negative wording) were transformed so that they shared the same sign and represented by the factor *wording*. For example, difficulty remained unchanged while easiness was multiplied by-1.

Subsequently, linear models were created for each attribute in the following format:

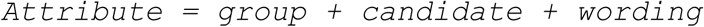

However, the *group* factor was not significant in any of the attribute models and was thus removed. The analysis was followed by pairwise contrasts between UACT test candidates using Bonferroni-corrected t-tests for comparison. Note that we use a linear model here with no random effects because of the standardization.

## Acknowledgements

This work was supported by Innovation Fund Denmark Grand Solutions 9090-00089B and the William Demant Foundation and carried out at the Technical University of Denmark.

We thank T. Dau and R.S. Sørensen for their support, the participants for their insights, time and effort, and L.B. Simonsen for her contributions to the custom procedure. The reverse digit span was originally implemented by J. Hjortkjaer and J. Märcher-Rørsted. We want to acknowledge the discussion among the UAud project partners and several colleagues from Demant A/S and DTU who tried out earlier versions of the tests.

## Author Contributions

RS-L: Conceptualization, Methodology, Software, Validation, Formal analysis, Investigation, Resources, Data Curation, Writing – Original Draft, Writing – Review & Editing, Visualization.

JZ: Conceptualization, Writing– Review & Editing, Investigation.

SL: Conceptualization, Writing – Review & Editing, Supervision, Project administration, Funding acquisition, Investigation, Methodology.

## Artificial Intelligence statement

During the preparation of this work, the authors used Gemini (version 2.5 Pro) and Mistral (version Nemo Instruct) to enhance readability and correct grammatical and typographical errors. The authors reviewed and edited all AI-generated suggestions and take full responsibility for the final content of the manuscript.

## Data availability

The data and code used in the analyses of this manuscript is available in the Zenodo repository https://doi.org/10.5281/zenodo.15730189. Most additional data can be made available on request.

## References

1. Haile, L. M. et al. Hearing loss prevalence and years lived with disability, 1990–2019: findings from the Global Burden of Disease Study 2019. The Lancet 397, 996–1009, (2021).

2. Shen, Y. et al. Global, regional, and national burden of hearing loss from 1990 to 2021: findings from the 2021 global burden of disease study. Ann. Med. 57, 2527367 (2025).

3. Clark, J. L. & Swanepoel, D. W. The World Report on Hearing – a new era for global hearing care. Int. J. Audiol. 60, 161–161 (2021).

4. Garuccio, J., Ukert, B., Arnold, M., Phillips, S. & Pesko, M. F. Using Supply and Demand to Identify Shortages in the Hearing Health Care Professional Workforce. JAMA Otolaryngol. Neck Surg. 151, 868 (2025).

5. Dillard, L. K. et al. Service delivery approaches related to hearing aids in low-and middle-income countries or resource-limited settings: A systematic scoping review. *PLOS Glob*. Public Health 4, e0002823 (2024).

6. Margolis, R. H. & Morgan, D. E. Automated Pure-Tone Audiometry: An Analysis of Capacity, Need, and Benefit. Am. J. Audiol. 17, 109–113 (2008).

7. Windmill, I. M. & Freeman, B. A. Demand for Audiology Services: 30-Yr Projections and Impact on Academic Programs. J. Am. Acad. Audiol. 24, 407–416 (2013).

8. Parmar, B. J., Rajasingam, S. L., Bizley, J. K. & Vickers, D. A. Factors Affecting the Use of Speech Testing in Adult Audiology. Am. J. Audiol. 31, 528–540 (2022).

9. Sidiras, C. User-Operated Audiometry Project (UAud) – Introducing an Automated User-Operated System for Audiometric Testing Into Everyday Clinic Practice. Front. Digit. Health 3, 7 (2021).

10. Gates, G. A. & Mills, J. H. Presbycusis. The Lancet 366, 1111–1120 (2005).

11. Houtgast, T. & Festen, J. M. On the auditory and cognitive functions that may explain an individual’s elevation of the speech reception threshold in noise. Int. J. Audiol. 47, 287–295 (2008).

12. Rönnberg, J. et al. Hearing impairment, cognition and speech understanding: exploratory factor analyses of a comprehensive test battery for a group of hearing aid users, the n200 study. Int. J. Audiol. 55, 623–642 (2016).

13. Schoof, T. & Rosen, S. The role of auditory and cognitive factors in understanding speech in noise by normal-hearing older listeners. Front. Aging Neurosci. 6, (2014).

14. Livingston, G. et al. Dementia prevention, intervention, and care: 2020 report of the Lancet Commission. The Lancet 396, 413–446 (2020).

15. Cantuaria, M. L. et al. Hearing Loss, Hearing Aid Use, and Risk of Dementia in Older Adults. JAMA Otolaryngol. Neck Surg. 150, 157 (2024).

16. Mata, R., Schooler, L. J. & Rieskamp, J. The aging decision maker: Cognitive aging and the adaptive selection of decision strategies. Psychol. Aging 22, 796–810 (2007).

17. Contreras-Somoza, L. M. et al. Usability and User Experience of Cognitive Intervention Technologies for Elderly People With MCI or Dementia: A Systematic Review. Front. Psychol. 12, 636116 (2021).

18. Zhang, Y., Zhu, X., Yang, S., Wong, A. K. C. & Chen, X. Digital Health Technologies Applied in Patients With Early Cognitive Change: Scoping Review. J. Med. Internet Res. 27, e82881 (2025).

19. Usability of a hearing test mobile app across generations | PLOS One. https://journals.plos.org/plosone/article?id=10.1371/journal.pone.0327726.

20. Grant, K. W., Walden, B. E., Summers, V. & Leek, M. R. Introduction: Auditory Models of Suprathreshold Distortion in Persons with Impaired Hearing. J. Am. Acad. Audiol. 24, 254–257 (2013).

21. Plomp, R. Auditory handicap of hearing impairment and the limited benefit of hearing aids. J. Acoust. Soc. Am. 63, 533–549 (1978).

22. Keidser, G. et al. The Quest for Ecological Validity in Hearing Science: What It Is, Why It Matters, and How to Advance It. Ear Hear. 41, 5S (2020).

23. Akeroyd, M. A. et al. International Collegium of Rehabilitative Audiology (ICRA) recommendations for the construction of multilingual speech tests: ICRA Working Group on Multilingual Speech Tests. Int. J. Audiol. 54, 17–22 (2015).

24. The Influence of Cochlear Mechanical Dysfunction, Temporal Processing Deficits, and Age on the Intelligibility of Audible Speech in Noise for Hearing-Impaired Listeners - Peter T. Johannesen, Patricia Pérez-González, Sridhar Kalluri, José L. Blanco, Enrique A. Lopez-Poveda, 2016. https://journals.sagepub.com/doi/full/10.1177/2331216516641055.

25. Strelcyk, O. & Dau, T. Relations between frequency selectivity, temporal fine-structure processing, and speech reception in impaired hearinga). J. Acoust. Soc. Am. 125, 3328–3345 (2009).

26. Bernstein, J. G. W. et al. Spectrotemporal Modulation Sensitivity as a Predictor of Speech Intelligibility for Hearing-Impaired Listeners. J. Am. Acad. Audiol. 24, 293–306 (2013).

27. Chi, T., Gao, Y., Guyton, M. C., Ru, P. & Shamma, S. Spectro-temporal modulation transfer functions and speech intelligibility. J. Acoust. Soc. Am. 106, 2719–2732 (1999).

28. Zaar, J., Simonsen, L. B., Dau, T. & Laugesen, S. Toward a clinically viable spectro-temporal modulation test for predicting supra-threshold speech reception in hearing-impaired listeners. Hear. Res. 427, 108650 (2023).

29. Levitt, H. Transformed up-down methods in psychoacoustics. J. Acoust. Soc. Am. 49, Suppl 2:467+ (1971).

30. Pelli, D. G. & Bex, P. Measuring contrast sensitivity. Vision Res. 90, 10–14 (2013).

31. Dorr, M. et al. Next-generation vision testing: the quick CSF. Curr. Dir. Biomed. Eng. 1, 131–134 (2015).

32. Mehraei, G., Gallun, F. J., Leek, M. R. & Bernstein, J. G. W. Spectrotemporal modulation sensitivity for hearing-impaired listeners: Dependence on carrier center frequency and the relationship to speech intelligibility. J. Acoust. Soc. Am. 136, 301–316 (2014).

33. Bernstein, J. G. W. et al. Spectrotemporal Modulation Sensitivity as a Predictor of Speech-Reception Performance in Noise With Hearing Aids. Trends Hear. 20, (2016).

34. Zaar, J., Simonsen, L. B. & Laugesen, S. A spectro-temporal modulation test for predicting speech reception in hearing-impaired listeners with hearing aids. Hear. Res. 443, 108949 (2024).

4. Zaar, J. / Simonsen, L. B., Sanchez-Lopez, R. & Laugesen, S. The Audible Contrast Threshold (ACT) test: A clinical spectro-temporal modulation detection test. Hear. Res. 453, 109103 (2024).

36. Margolis, R. H., Glasberg, B. R., Creeke, S. & Moore, B. C. J. AMTAS®: Automated method for testing auditory sensitivity: Validation studies. Int. J. Audiol. 49, 185–194 (2010).

37. Larusdottir, M. K., Gulliksen, J. & Hallberg, N. RAMES – Framework supporting user centred evaluation in research and practice. Behav. Inf. Technol. 38, 132–149 (2019).

29. ISO/IEC 25022: Systems and software engineering — Systems and software Quality Requirements and Evaluation (SQuaRE) - Measurement of quality in use (2016)

39. Blackburn, H. L. & Benton, A. L. Revised administration and scoring of the Digit Span Test. J. Consult. Psychol. 21, 139–143 (1957).

40. Fuglsang, S. A., Märcher-Rørsted, J., Dau, T. & Hjortkjær, J. Effects of Sensorineural Hearing Loss on Cortical Synchronization to Competing Speech during Selective Attention. J. Neurosci. 40, 2562–2572 (2020).

41. Ericsson, K. A. & Simon, H. A. How to Study Thinking in Everyday Life: Contrasting Think-Aloud Protocols With Descriptions and Explanations of Thinking. Mind Cult. Act. 5, 178–186 (1998).

42. Carhart, R. & Jerger, J. F. Preferred Method For Clinical Determination Of Pure-Tone Thresholds. J. Speech Hear. Disord. 24, 330–345 (1959).

43. Jebb, A. T., Ng, V. & Tay, L. A Review of Key Likert Scale Development Advances: 1995–2019. Front. Psychol. 12, (2021).

44. Koo, T. K. & Li, M. Y. A Guideline of Selecting and Reporting Intraclass Correlation Coefficients for Reliability Research. J. Chiropr. Med. 15, 155–163 (2016).

45. Bartlett, J. W. & Frost, C. Reliability, repeatability and reproducibility: analysis of measurement errors in continuous variables. Ultrasound Obstet. Gynecol. 31, 466–475 (2008).

46. Bland, J. M. & Altman, D. G. Statistical methods for assessing agreement between two methods of clinical measurement. Int. J. Nurs. Stud. 6 (2010).

47. Giavarina, D. Understanding Bland Altman analysis. *Biochem*. Medica 25, 141–151 (2015).

48. Dorr, M. et al. New Precision Metrics for Contrast Sensitivity Testing. IEEE J. Biomed. Health Inform. 22, 919–925 (2018).

49. Sørensen, C. B., Pedersen, C., Pedersen, E. R., Schmidt, J. H. & Nielsen, J. Observed issues and their prevalence when patients conduct user-operated audiometry: International Symposium on Auditory and Audiological Research. in (2023).

50. Aronoff, J. M. & Landsberger, D. M. The development of a modified spectral ripple test. J. Acoust. Soc. Am. 134, EL217–EL222 (2013).

51. Landsberger, D. M., Dwyer, R. T., Stupak, N. & Gifford, R. H. Validating a Quick Spectral Modulation Detection Task. Ear Hear. 40, 1478–1480 (2019).

52. Lelo De Larrea-Mancera, E. S., et al. Portable Automated Rapid Testing (PART) for auditory assessment: Validation in a young adult normal-hearing population. J. Acoust. Soc. Am. 148, 1831–1851 (2020).

53. Landsberger, D. M., Dwyer, R. T., Stupak, N. & Gifford, R. H. Validating a Quick Spectral Modulation Detection Task. Ear Hear. 40, 1478 (2019).

54. Lelo de Larrea-Mancera, E. S., et al. At-Home Auditory Assessment Using Portable Automated Rapid Testing (PART) to Understand Self-Reported Hearing Difficulties. Trends Hear. 29, 23312165251397373 (2025).

55. Stavropoulos, T. A. et al. Exponential spectro-temporal modulation generation. J. Acoust. Soc. Am. 149, 1434–1443 (2021).

56. Lelo de Larrea-Mancera, E. S., et al. Portable Automated Rapid Testing (PART) for auditory assessment: Validation in a young adult normal-hearing population. J. Acoust. Soc. Am. 148, 1831–1851 (2020).

57. Lelo de Larrea-Mancera, E. S., et al. Remote auditory assessment using Portable Automated Rapid Testing (PART) and participant-owned devicesa). J. Acoust. Soc. Am. 152, 807–819 (2022).

58. Lelo de Larrea-Mancera, E. S., et al. Validation of the adaptive scan method in the quest for time-efficient methods of testing auditory processes. Atten. Percept. Psychophys. 85, 2797–2810 (2023).

59. Shojaeemend, H. & Ayatollahi, H. Automated Audiometry: A Review of the Implementation and Evaluation Methods. Healthc. Inform. Res. 24, 263 (2018).

60. Eikelboom, R. H., Swanepoel, D. W., Motakef, S. & Upson, G. S. Clinical validation of the AMTAS automated audiometer. Int. J. Audiol. 52, 342–349 (2013).

61. Margolis, R. H., Killion, M. C., Bratt, G. W. & Saly, G. L. Validation of the Home Hearing Test^TM^. J. Am. Acad. Audiol. 27, 416–420 (2016).

62. Pedersen, C. et al. Comparison of hearing-aid effectiveness based on user-operated versus traditional audiometry: a randomised clinical trial. Int. J. Audiol. 1–8 (2024) doi:10.1080/14992027.2024.2434897.

63. Voutilainen, A., Pitkäaho, T., Kvist, T. & Vehviläinen-Julkunen, K. How to ask about patient satisfaction? The visual analogue scale is less vulnerable to confounding factors and ceiling effect than a symmetric Likert scale. J. Adv. Nurs. 72, 946–957 (2016).

64. Sung, Y.-T. & Wu, J.-S. The Visual Analogue Scale for Rating, Ranking and Paired-Comparison (VAS-RRP): A new technique for psychological measurement. Behav. Res. Methods 50, 1694–1715 (2018).

65. García-Pérez, M. A. & Alcalá-Quintana, R. Accuracy and precision of responses to visual analog scales: Inter-and intra-individual variability. Behav. Res. Methods 55, 4369–4381 (2023).

66. Kersten, P., White, P. J. & Tennant, A. Is the Pain Visual Analogue Scale Linear and Responsive to Change? An Exploration Using Rasch Analysis. PLOS ONE 9, e99485 (2014).

67. Nasreddine, Z. S., et al. The Montreal Cognitive Assessment, MoCA: a brief screening tool for mild cognitive impairment. J. Am. Geriatr. Soc. 53, 695–699 (2005).

68. Diniz, B. S. O., Yassuda, M. S., Nunes, P. V., Radanovic, M. & Forlenza, O. V. Mini-mental State Examination performance in mild cognitive impairment subtypes. Int. Psychogeriatr. 19, 647–656 (2007).

69. Poling, G. L., Kunnel, T. J. & Dhar, S. Comparing the Accuracy and Speed of Manual and Tracking Methods of Measuring Hearing Thresholds: Ear Hear. 37, e336–e340 (2016).

71. Mahomed, F., Swanepoel, D. W., Eikelboom, R. H. & Soer, M. Validity of Automated Threshold Audiometry: A Systematic Review and Meta-Analysis. Ear Hear. 34, 745–752 (2013).

72. Nielsen, J. & Landauer, T. K. A mathematical model of the finding of usability problems. in Proceedings of the SIGCHI conference on Human factors in computing systems - CHI’93 206–213 (ACM Press, Amsterdam, The Netherlands, 1993). doi:10.1145/169059.169166.

73. MIkhalevskaya, M. B. & FInkel’, N. V. The Influence of Feedback on the Detectability of a Signal. Sov. Psychol. 25, 50–60 (1986).

74. Hoglund, E. M. & Feth, L. L. Bayesian analysis of the impact of incomplete feedback on response bias. in 050006 (Sydney, Australia, 2023). doi:10.1121/2.0001851.

75. Suh, M. J. et al. Improving Accuracy and Reliability of Hearing Tests: An Exploration of International Standards. J. Audiol. Otol. 27, 169–180 (2023).

76. Remus, J. J. & Collins, L. M. Comparison of adaptive psychometric procedures motivated by the Theory of Optimal Experiments: Simulated and experimental results. J. Acoust. Soc. Am. 123, 315–326 (2008).

77. Brand, T. & Kollmeier, B. Efficient adaptive procedures for threshold and concurrent slope estimates for psychophysics and speech intelligibility tests. J. Acoust. Soc. Am. 111, 2801–2810 (2002).

78. Zabalbeascoa, P. The nature of the audiovisual text and its parameters. in The Didactics of Audiovisual Translation vol. 77 21–37 (John Benjamins Publishing, 2008).

79. McGraw, K. O. & Wong, S. P. Forming inferences about some intraclass correlation coefficients. Psychol. Methods 1, 30–46 (1996).

80. Dorr, M. et al. New Precision Metrics for Contrast Sensitivity Testing. IEEE J. Biomed. Health Inform. 22, 919–925 (2018).

81. Margolis, R. H., Saly, G. L., Le, C. & Laurence, J. Qualind: A method for assessing the accuracy of automated tests. J. Am. Acad. Audiol. 18, 78–89 (2007).

